# IMPACT OF WEEKNIGHT AND WEEKEND CURFEWS USING MOBILITY DATA: A CASE STUDY OF BENGALURU URBAN

**DOI:** 10.1101/2022.01.26.22269903

**Authors:** Aniruddha Adiga, Siva Athreya, Madhav Marathe, Jagadish Midthala, Nihesh Rathod, Rajesh Sundaresan, Srinivasan Venkataramanan, Sarath Yasodharan

## Abstract

Karnataka imposed weeknight and weekend curfews to mitigate the spread of the Omicron variant of SARS-CoV-2. We attempt to assess the impact of curfew using community mobility reports published by Google. Then, we quantify the impact of such restrictions via a simulation study. The pattern of weeknight and weekend curfew, followed by relaxations during the weekdays, seems, at best, to slow and delay the Omicron spread. The simulation outcomes suggest that Omicron eventually spreads and affects nearly as much of the population as it would have without the restrictions. Further, if Karnataka cases trajectory follows the South African Omicron wave trend and the hospitalisation is similar to that observed in well-vaccinated countries (2% of the confirmed cases), then the healthcare requirement is likely within the capacity of Bengaluru Urban when the caseload peaks, with or without the mobility restrictions. On the other hand, if Karnataka cases trajectory follows both the South African Omicron wave trend and the hospitalisation requirement observed there (6.9%), then the healthcare capacity may be exceeded at peak, with or without the mobility restrictions.

## Introduction

Since the last week of December 2021, India has seen a significant resurgence in COVID-19 cases due to the Omicron variant (World Health Organization, 2021). Given the steep increase in cases in other well-vaccinated parts of the world, mainly Europe and the United States, and anticipating a similar rise in India, the state of Karnataka imposed weeknight and weekend curfews starting the night of 07 Jan 2022 to mitigate the spread. The purpose of this modelling study is to quantify the public health benefit of such intermittent curfews. As a case study, we focus on the impact of the curfew-related restrictions on the Bengaluru Urban district. This urbanised district has the largest population in Karnataka at an estimated 13.6 million and has been the most impacted in all the waves.

## Methods

### An attempt to assess the impact of curfew via mobility data

We focus attention on the reduction in mobility due to the weeknight and weekend curfew. Since the start of the COVID-19 pandemic, Google Inc. has been publishing community mobility reports (Google Inc., 2022) that indicate the level of activity in various interaction spaces. These community mobility reports quantify the reduction (or increase) in the mobility on a given day with respect to the normal baseline mobility for that day of the week; the benchmark or baseline is a certain specified pre-pandemic period (Google Inc., 2022). We extracted information from the data and report of 15 Jan 2022 (Google Inc., 2022) and plotted the mobility factors for each day of the week in Figure 2 of the Appendix. Table 1 below summarises the mobility level in the two weeks of January 2022 with respect to the average level in December 2021.

**Table 1.**
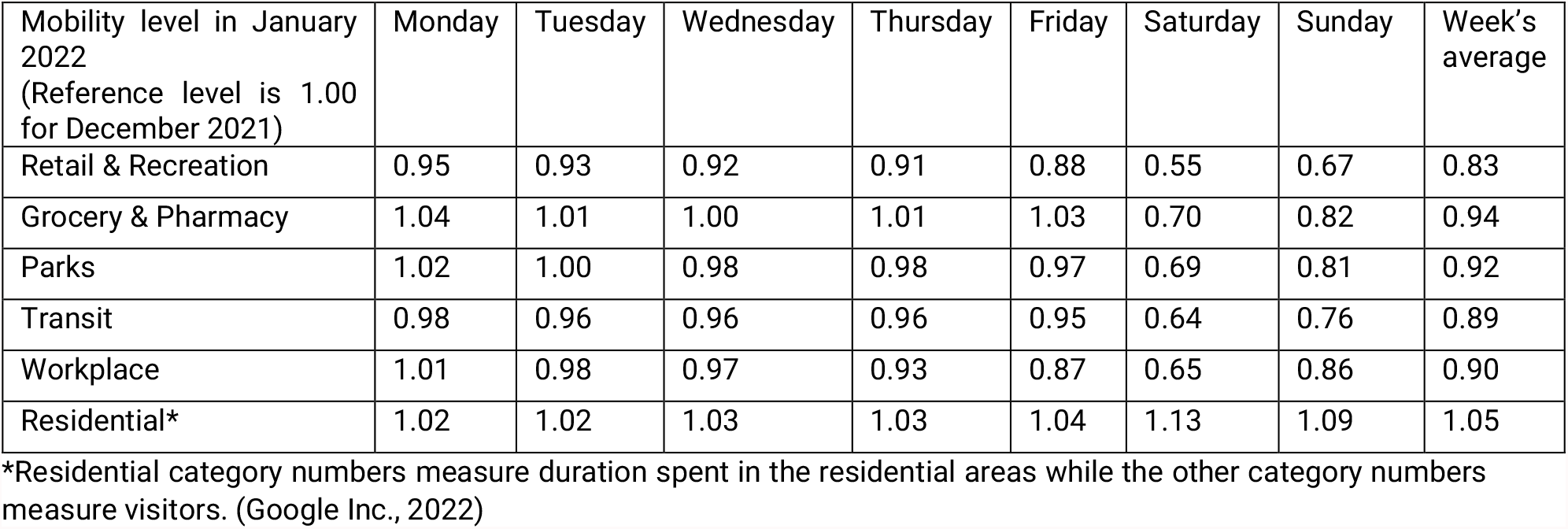
Summary of Google Community Mobility Report (Google Inc., 2022). Each number is a factor by which mobility has decreased/increased on that day of the week in the first two weeks of January 2022 with respect to the average for the corresponding day of the week in December 2021 taken as the reference period. A value smaller than 1 implies a reduction in mobility. The residential category numbers measure the duration spent in the area, whereas the other category numbers measure visitors.

| Mobility level in January 2022<br>(Reference level is 1.00<br>for December 2021) | Monday | Tuesday | Wednesday | Thursday | Friday | Saturday | Sunday | Week's<br>average |
| --- | --- | --- | --- | --- | --- | --- | --- | --- |
| Retail & Recreation | 0.95 | 0.93 | 0.92 | 0.91 | 0.88 | 0.55 | 0.67 | 0.83 |
| Grocery & Pharmacy | 1.04 | 1.01 | 1.00 | 1.01 | 1.03 | 0.70 | 0.82 | 0.94 |
| Parks | 1.02 | 1.00 | 0.98 | 0.98 | 0.97 | 0.69 | 0.81 | 0.92 |
| Transit | 0.98 | 0.96 | 0.96 | 0.96 | 0.95 | 0.64 | 0.76 | 0.89 |
| Workplace | 1.01 | 0.98 | 0.97 | 0.93 | 0.87 | 0.65 | 0.86 | 0.90 |
| Residential* | 1.02 | 1.02 | 1.03 | 1.03 | 1.04 | 1.13 | 1.09 | 1.05 |

\*Residential category numbers measure duration spent in the residential areas while the other category numbers measure visitors. (Google Inc., 2022)

The data in Table 1 uses aggregated and anonymised sets of data from smartphone users who have turned on the Location History setting; this is set to ‘off’ by default (Google Inc., 2022). Consequently, we recognise that this will have a bias in assessing overall mobility, which will carry to our assessment of the curfews. With this caveat, we make the following observations from Table 1.

1. In the two weeks of January starting 01 Jan 2022, the first week does not have mobility restrictions whereas the second week has mobility restrictions.
2. There is a marginal reduction in the retail & recreation, transit, and workplace category mobilities during the weekdays, accompanied by a marginal increase in the grocery & pharmacy category mobility and the time spent in the residential areas.
3. Weekend mobility reduces in all non-residential categories.
4. Duration of time spent in the residential areas increases by about 9-13% over the weekend.
5. The above table suggests an approximately 10% reduction in mobility over the two weeks, taking an average across all categories except the residential category; see the last column in Table 1.

Therefore, in view of the first point, we assume a 20% reduction in mobility in this study on account of the mobility restrictions. This reduction may not be entirely due to the weeknight/weekend curfews since there has been a significant behavioural adaptation to the Omicron spread arising from the increased information and communication campaigns and the greater public awareness. The mobility charts in Figure 2 of the Appendix indicate similar mobility reductions in April 2021, during the second wave, well before the Karnataka lockdown on 26 Apr 2021. Given this, we also provide estimates for the assumptions of 5%, 10%, and 15% reduction in mobility.

### Model

The IISc-ISI team, in collaboration with the University of Virginia’s Biocomplexity Institute, developed a mean-field model to understand the district-wise COVID-19 spread during the April 2021 wave. We studied the impact of antibody waning and prioritising vaccination across the districts of Karnataka (Adiga, et al., 2021). Recently this model has been adapted to provide the Omicron projections (ISI-IISc-Team, 2022). A brief description of the adaptation follows. We use an SEIR model with antibody waning and vaccination (with vaccination effectiveness taken into account). We use the April 2021 wave, when the Delta variant was predominant, as the base case in each district. We then estimate the Omicron-to-Delta transmissibility factor from a study of the South Africa data (Department of Health, Republic of South Africa, 2022). We then assume that the biological factors leading to the increased transmissibility of the Omicron variant over the Delta variant are the same in South Africa and India and apply that same factor to estimate the transmissibility of Omicron in Karnataka. The population level of immunity to the Omicron variant is uncertain. We therefore use three levels of population-level susceptibilities in the study: 30%, 60%, and a worst-case 100% susceptibility. The Omicron wave then starts as a new infection spread for each of these three levels of susceptibilities. The projection of cases, updated on 19 Jan 2022, is given below for the reference case of no restrictions (i.e., no curfew imposed).

The Figure 1 estimates are higher than the projections provided on 17 Jan 2022 in (ISI-IISc-Team, 2022) to account for an increased level of testing in the early part of January 2022 which likely leads to increased case ascertainment. Similarly, decreased level of testing leads to reduced case ascertainment. Furthermore, the large level of asymptomatic infection associated with the Omicron variant also leads to reduced case ascertainment. Holding the number of tests to be a constant into the future (18 Jan 2022 onwards), based on the average for the week ending 17 Jan 2022, the model projects a peak of about 52,348 cases around 23 Jan 2022 for the 30% susceptibility scenario and roughly double this number with a peak five days later for the 60% susceptibility scenario.

**Figure 1.**
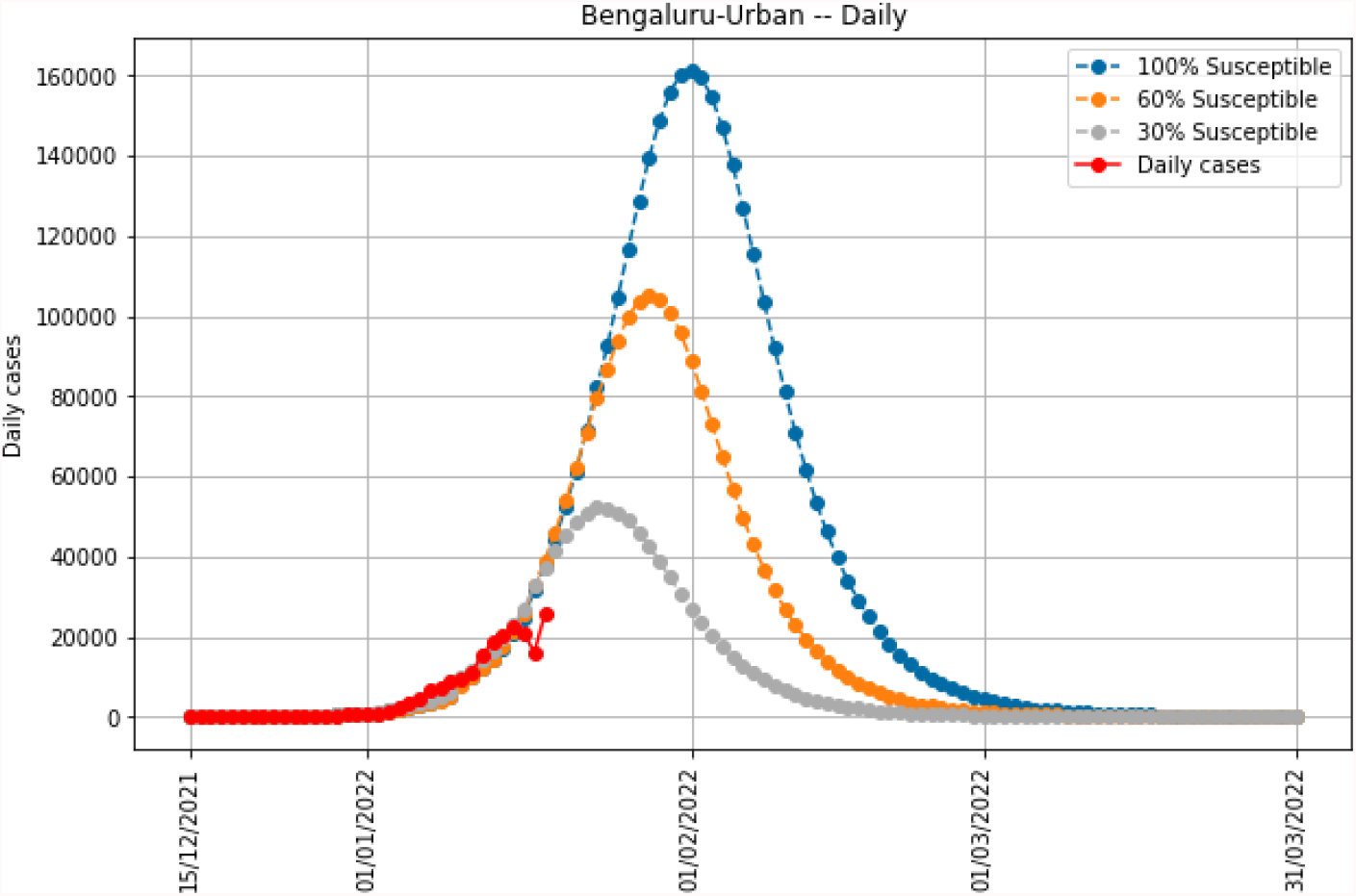
Daily cases trajectory in Bengaluru Urban. This is without any mobility restriction using the estimated contact rate until 31 Mar 2022.

### Simulation experiment design

We now describe the simulation experiments that we conducted on the model.

Based on a study of Table 1, we considered the following effective mobility factors for our case study: 95%, 90%, 85%, and 80%. We did not go below 80% because the increase in the residential area dwell times during curfews is likely to compensate to some extent the mobility reduction in the other areas. Further, as noted earlier, the reduction cannot be attributed to the curfew alone since there is a natural reduction trend due to increased awareness.

We assume that the imposed mobility restrictions result from the weeknight and weekend curfews, as was introduced in Karnataka. We then consider the following scenarios:

- Base case scenario: No mobility restrictions.
- Weekly mobility restrictions as described above until 31 Mar 2022.

We also compare scenarios that lift the mobility restrictions at various dates before 31 Mar 2022.

For understanding the impact in each scenario, we compare the following quantities.

- The cumulative number of cases from 15 Dec 2021 to 31 Jan 2022.
- The cumulative number of cases from 15 Dec 2021 to 28 Feb 2022.
- The cumulative number of cases from 15 Dec 2021 to 31 Mar 2022.
- The peak number of cases during this wave.
- The hospital beds requirement at when the caseload peaks.
- The ICU beds-with-ventilator requirement when the caseload peaks.

## Results

1. We compare the base scenario with the other scenario when mobility is restricted until 31 Mar 2022. To do this, we specifically compare cases at the end of January, February, and March 2022. The reductions in the number of cases due to the mobility restrictions are substantial at the end of January 2022 under the 30% and 60% susceptibility assumptions. However, the smaller reductions at the end of February 2022 and at the end of March 2022 indicate that the infections eventually rise and come close to the level of the base case scenario of no restrictions. A standard mathematical analysis suggests that the cumulative cases reach a fixed steady-state that depends only on the contact rate and recovery rate, even if contact rates were reduced through curfews but only temporarily. The simulation results point to something more significant in the dynamics. For the specific parameters, despite mobility reduction until 31 Mar 2022, the cumulative infections are close to catching up as early as 28 Feb 2022, which one might argue is due to the increased transmissibility of the Omicron variant. Therefore, the benefit of the mobility restrictions is to ‘flatten the (cases) curve’ and reduce the peak.
2. One of the crucial benefits of flattening the curve is reducing the daily demand for hospital beds. We now quantify this benefit. We look at the peak cases for the various scenarios and estimate the number of hospital beds required. It may be noted for comparison purposes that the numbers of available hospital beds and ICU beds with ventilators (capacities) in Bengaluru Urban are 7917 and 450, respectively (Bruhat Bengaluru Mahanagara Palike, 2022) as on 19 Jan 2022. Efforts are ongoing to increase the capacities. Going by the data from South Africa (Department of Health, Republic of South Africa, 2022), conservatively, we assume 6.9% of the cases will need hospital beds. In this case, the requirements at the peak in Bengaluru Urban are summarised in Table 3. (The estimate of ICU beds with ventilators assumes that 5% of the hospitalised cases will need this support.) Similarly, we consider data from the UK and assume that only 2% of the cases will need hospital beds, 5% of the hospitalised patients will need ICU beds with ventilators, and provide the modified requirements in the following table.
3. The actual mobility may be higher than the 80% assumed above. We consider alternative mobility scenarios at 85%, 90%, and 95% of the December 2021 mobility and simulate these scenarios to provide the following estimates (Table 5) of hospital beds. The forecast of ICU Beds with ventilators is calculated as 5% of the indicated forecast of hospital beds. We also consider a scenario where we remove the restriction on 21 Jan 2022 but then bring back the restrictions after 05 Feb 2022. The table entries are self-explanatory.

## Discussion

We first describe the limitations of our study. The analysis and hospitalisation forecast assumes that the Omicron variant of SARS-CoV-2 is the dominant variant. However, if the Delta variant is still prevalent, hospitalisation and the severity estimations may differ from those for the Omicron variant. Regular and timely sequencing data will tell us whether the Delta variant is still in circulation or not. Further, our study focused only on cases and associated estimates of required hospital beds. In particular, we have not examined several other important factors, e.g., workforce shortages, overcrowding events in enclosed spaces, etc. Moreover, our model has a homogenised contact rate for the entire week, and the impact of the curfew is modelled as a reduction in the weekly contact rate. Finally, as already highlighted, our assessment of the curfew is based on Google’s published community mobility reports which may suffer from an inherent sampling bias.

With these caveats highlighted, we now discuss the outcomes of our simulation case study for Bengaluru Urban.

1. The pattern of weeknight and weekend curfew, followed by relaxations during the weekday, seems, at best, to slow and delay the Omicron spread. The simulation outcomes suggest that Omicron eventually spreads across the population. The *cumulative reduction* at the end of March 2022 is 10,113 for the 30% susceptibility assumption and 19,889 for the 60% susceptibility assumption. These forecasts are out of the projected cumulative cases of 9,94,228 (30%) and 20,12,019 (60%), respectively, at the end of March 2022 (Table 2).
2. When mobility restrictions are in force until March 2022, the reduction in the peak cases is 9,399 (30% susceptibility) and 18,326 (60% susceptibility), see Table 3. Since the peak is expected in the third week of January 2022, the usefulness of the restrictions beyond the peak is limited.
3. The required number of hospital beds, when the caseload peaks, is 7328 without mobility restrictions (at 30% susceptibility, 2% hospitalisation, Table 4). The forecast of hospital beds does not exceed Bengaluru’s capacity of 7917. However, at (2% hospitalisation, 60% susceptibility assumption) and at 6.9% hospitalisation and either 30% or 60% susceptibility assumptions, the hospital bed capacity is exceeded at peak even with the mobility restrictions. Given that mobility is allowed during the weekdays, the impact of the weeknight and weekend curfew is therefore limited.

**Table 2.** Comparison of cases.

| Scenario | 31 Jan (30% susc.) | 28 Feb (30% susc.) | 31 Mar (30% susc.) | 31 Jan (60% susc.) | 28 Feb (60% susc.) | 31 Mar (60% susc.) |
| --- | --- | --- | --- | --- | --- | --- |
| Base case (no mobility restrictions) | 806131 | 991616 | 994228 | 1308304 | 2003132 | 2012019 |
| 80% mobility till end of March 2022 | 684290 | 977499 | 984115 | 960802 | 1966777 | 1992130 |
| Reduction in the cumulative number of cases on the indicated date | 121841 | 14117 | 10113 | 347502 | 36355 | 19889 |

**Table 3.**
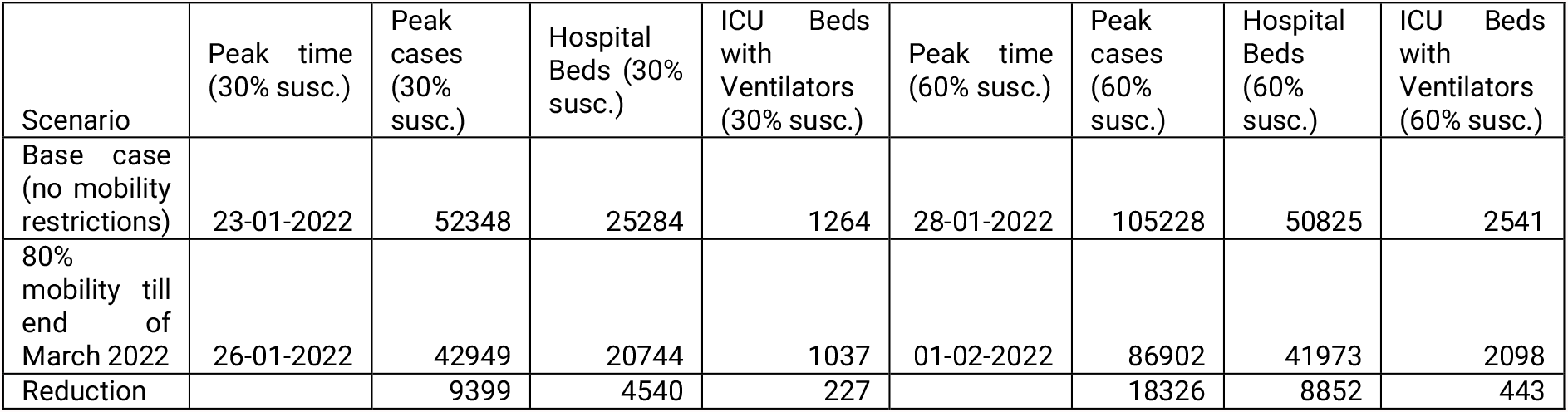
Hospital bed requirement at peak assuming 6.9% of the cases need hospitalisation (which is derived from South Africa Omicron wave data)

| Scenario | Peak time (30% susc.) | Peak cases (30% susc.) | Hospital Beds (30% susc.) | ICU Beds with Ventilators (30% susc.) | Peak time (60% susc.) | Peak cases (60% susc.) | Hospital Beds (60% susc.) | ICU Beds with Ventilators (60% susc.) |
| --- | --- | --- | --- | --- | --- | --- | --- | --- |
| Base case (no mobility restrictions) | 23-01-2022 | 52348 | 25284 | 1264 | 28-01-2022 | 105228 | 50825 | 2541 |
| 80% mobility till end of March 2022 | 26-01-2022 | 42949 | 20744 | 1037 | 01-02-2022 | 86902 | 41973 | 2098 |
| Reduction |  | 9399 | 4540 | 227 |  | 18326 | 8852 | 443 |

**Table 4.** Hospital bed requirement at peak assuming 2% of the cases need hospitalisation (based on UK Omicron wave data on 17/01/2022)

| Scenario | Peak time<br>(30% susc.) | Peak cases<br>(30% susc.) | Hospital<br>Beds (30%<br>susc.) | ICU Beds<br>with<br>Ventilators<br>(30% susc.) | Peak time<br>(60% susc.) | Peak cases<br>(60% susc.) | Hospital<br>Beds<br>(60%<br>susc.) | ICU Beds<br>with<br>Ventilators<br>(60% susc.) |
| --- | --- | --- | --- | --- | --- | --- | --- | --- |
| Base case<br>(no mobility<br>restrictions) | 23-01-2022 | 52348 | 7328 | 366 | 28-01-2022 | 105228 | 14731 | 736 |
| 80%<br>mobility till<br>end of<br>March 2022 | 26-01-2022 | 42949 | 6012 | 300 | 01-02-2022 | 86902 | 12166 | 608 |
| Reduction |  | 9399 | 1316 | 66 |  | 18326 | 2565 | 128 |

**Table 5.** Hospital beds at peak time for all mobility scenarios considered.

| Scenario | Peak time<br>(30% susc.) | Peak cases<br>(30% susc.) | Hospital<br>Beds at<br>6.9%<br>(30%<br>susc.) | Hospital<br>Beds at 2%<br>(30%<br>susc.) | Peak time<br>(60% susc.) | Peak cases<br>(60% susc.) | Hospital<br>Beds at<br>6.9% (60%<br>susc.) | Hospital<br>Beds at<br>2% (60%<br>susc.) |
| --- | --- | --- | --- | --- | --- | --- | --- | --- |
| Base case<br>scenario (full<br>mobility) | 23-01-2022 | 52348 | 25284 | 7328 | 28-01-2022 | 105228 | 50825 | 14731 |
| 95% mobility<br>until 21 Jan<br>2022 only. | 24-01-2022 | 50573 | 24426 | 7080 | 29-01-2022 | 104199 | 50328 | 14587 |
| 90% mobility<br>until 21 Jan<br>2022 only | 25-01-2022 | 49703 | 24006 | 6958 | 29-01-2022 | 103259 | 49874 | 14456 |
| 85% mobility<br>until 21 Jan<br>2022 only | 26-01-2022 | 48849 | 23594 | 6838 | 30-01-2022 | 102692 | 49600 | 14376 |
| 80% mobility<br>until 21 Jan<br>2022 only | 26-01-2022 | 48076 | 23220 | 6730 | 31-01-2022 | 101961 | 49247 | 14274 |
| 95% mobility<br>until 31 Jan<br>2022 only | 23-01-2022 | 49770 | 24038 | 6967 | 29-01-2022 | 101042 | 48803 | 14145 |
| 90% mobility<br>until 31 Jan<br>2022 only | 25-01-2022 | 47620 | 23000 | 6666 | 30-01-2022 | 96540 | 46628 | 13515 |
| 85% mobility<br>until 31 Jan<br>2022 only | 25-01-2022 | 45275 | 21867 | 6338 | 31-01-2022 | 91822 | 44350 | 12855 |
| 80% mobility<br>until 31 Jan<br>2022 only | 26-01-2022 | 42950 | 20744 | 6013 | 02-02-2022 | 89076 | 43023 | 12470 |
| 95% mobility<br>throughout | 23-01-2022 | 49771 | 24039 | 6967 | 29-01-2022 | 101041 | 48802 | 14145 |
| 90% mobility<br>throughout | 25-01-2022 | 47620 | 23000 | 6666 | 30-01-2022 | 96540 | 46628 | 13515 |
| 85% mobility throughout | 25-01-2022 | 45275 | 21867 | 6338 | 31-01-2022 | 91822 | 44350 | 12855 |
| 80% mobility throughout | 26-01-2022 | 42949 | 20744 | 6012 | 01-02-2022 | 86902 | 41973 | 12166 |
| 95% mobility until 21 Jan 2022, full mobility until 05 Feb 2022, and then 95% mobility thereafter | 24-01-2022 | 50573 | 24426 | 7080 | 29-01-2022 | 104199 | 50328 | 14587 |
| 90% mobility until 21 Jan 2022, full mobility until 05 Feb 2022, and then 90% mobility thereafter | 25-01-2022 | 49704 | 24007 | 6958 | 29-01-2022 | 103259 | 49874 | 14456 |
| 85% mobility until 21 Jan 2022, full mobility until 05 Feb 2022, and then 85% mobility thereafter | 26-01-2022 | 48849 | 23594 | 6838 | 30-01-2022 | 102691 | 49599 | 14376 |
| 80% mobility until 21 Jan 2022, full mobility until 05 Feb 2022, and then 80% mobility thereafter | 26-01-2022 | 48076 | 23220 | 6730 | 31-01-2022 | 101960 | 49246 | 14274 |

Reduced testing levels or increased home testing may bring the case counts down, resulting in an overestimation of the predicted cases. But estimates of hospitalisation and ICU beds with ventilators may yet be robust since individuals needing such care may eventually seek healthcare support.

Our study is yet another example of the vital role models can play in assisting policymakers in making more informed choices. However, a critical factor in enabling this is data availability. The *timely* availability of *statistically significant* variant estimates, giving the mix of the Delta and the Omicron variants in Bengaluru Urban, can lead to a better estimation of the required hospital beds. We need a sufficient number of positive samples to be sequenced in each district for statistical significance. Anonymised age, co-morbidities, and the vaccination or previous disease status data of confirmed cases can strengthen modelling and offer policymakers a greater variety of policy instruments. For example, these could result in more focused guidance on the timing of school and college openings, prioritisation of vaccination in multigenerational homes, etc. Finally, the methodology employed in our study will have wider applicability for other cities and districts.

## Data Availability

All data produced in the present work are contained in the manuscript

https://www.incovid19.org/

https://apps.bbmpgov.in/Covid19/en/media_pdf/Covid_Bengaluru_19_January_2022%20Bulletin-667.pdf

https://www.nicd.ac.za/wp-content/uploads/2022/01/NICD-COVID-19-Daily-Sentinel-Hospital-Surveillance-report-National-20220102.pdf

https://www.google.com/covid19/mobility/

https://www.gstatic.com/covid19/mobility/2022-01-15_IN_Karnataka_Mobility_Report_en-GB.pdf

## Acknowledgements

This work was partially supported by the Centre for Networked Intelligence, the Indian Statistical Institute’s CPDA, the NSF Expedition in Computing Grant CCF-1918656, the NSF RAPID CCF-2142997 and the IISc Institution of Eminence grants. Any opinions, findings, conclusions, or recommendations expressed in this material are those of the authors and do not necessarily reflect the views of the funding agencies.

## Appendix

**Figure 2.**
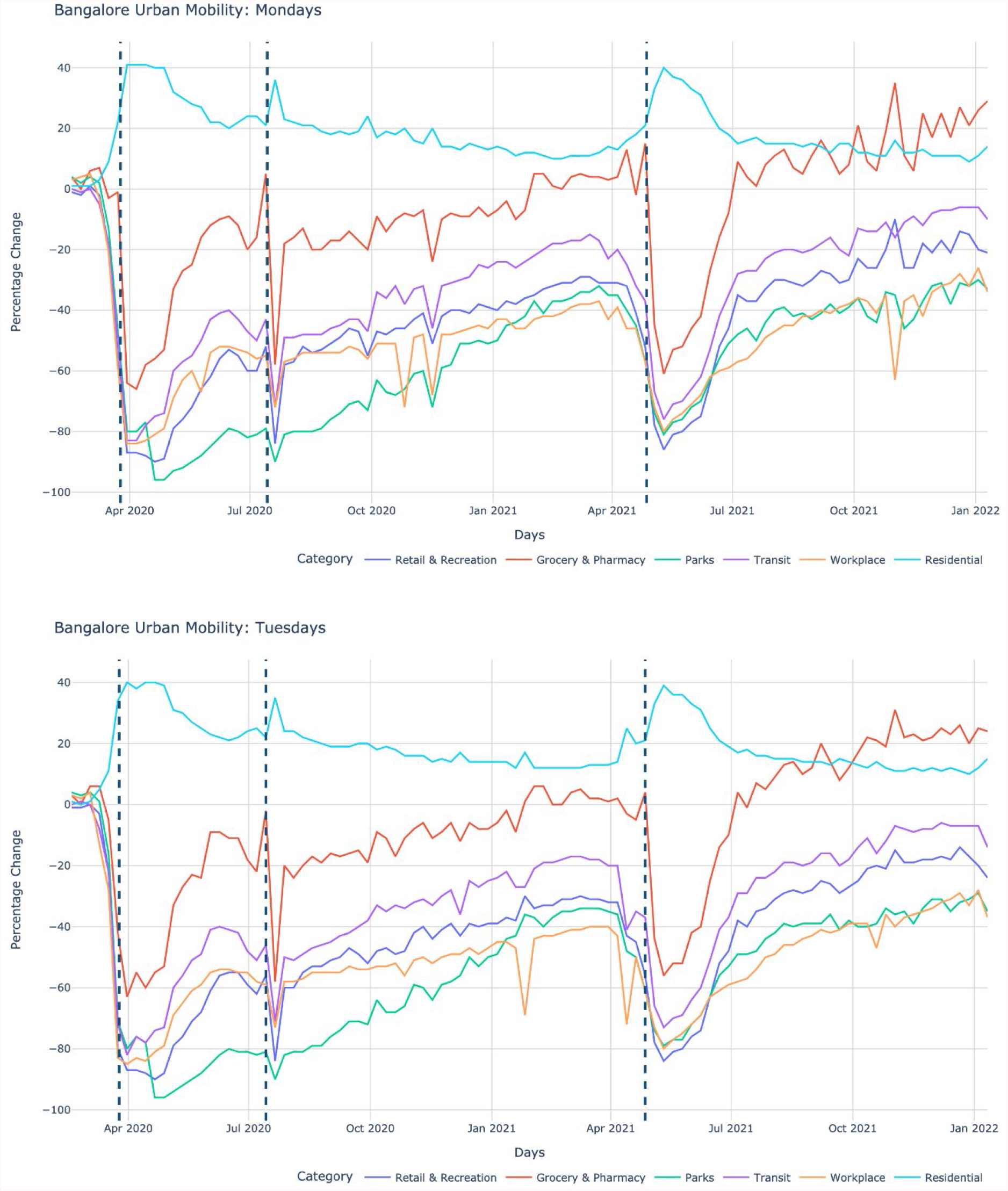
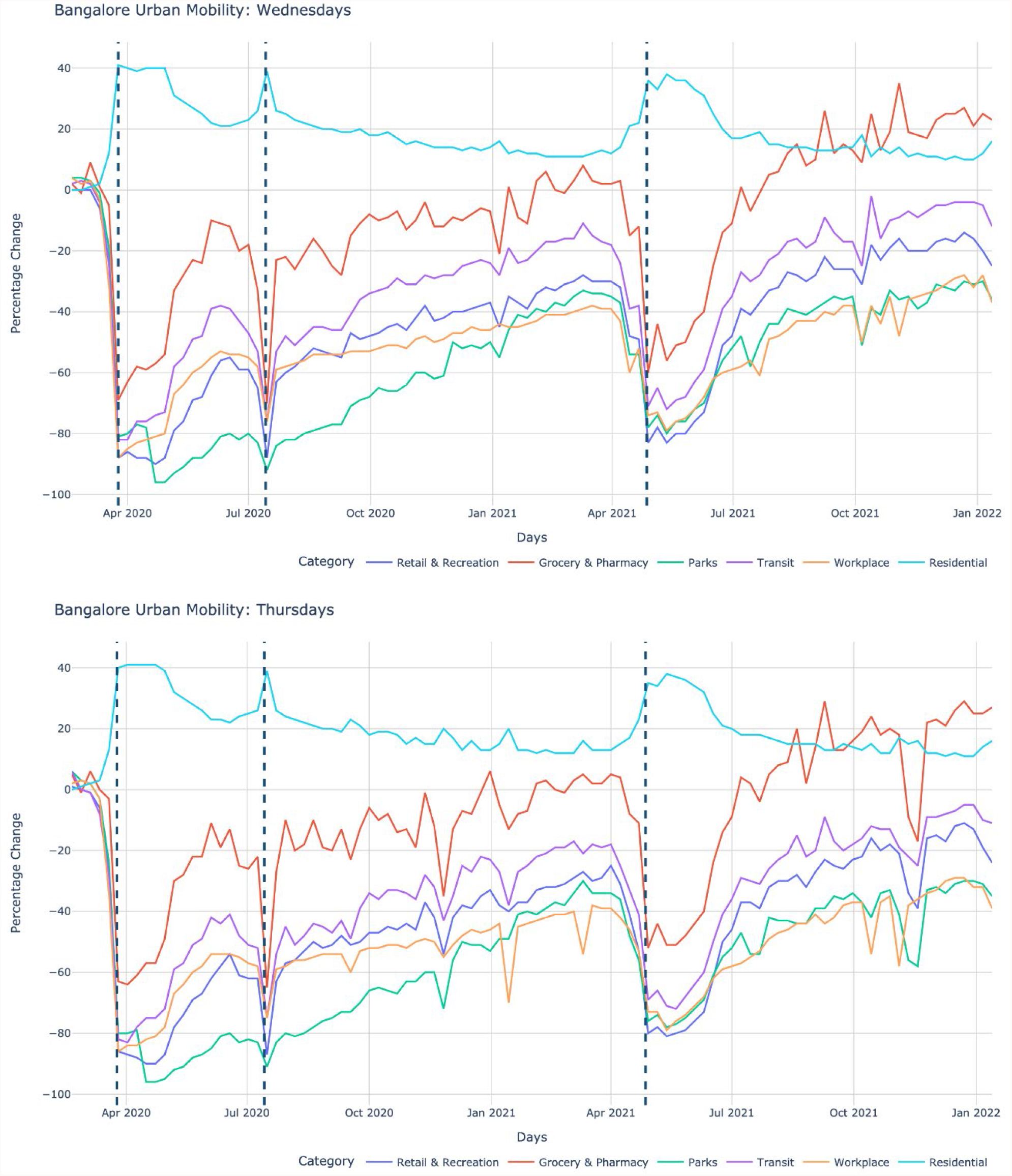
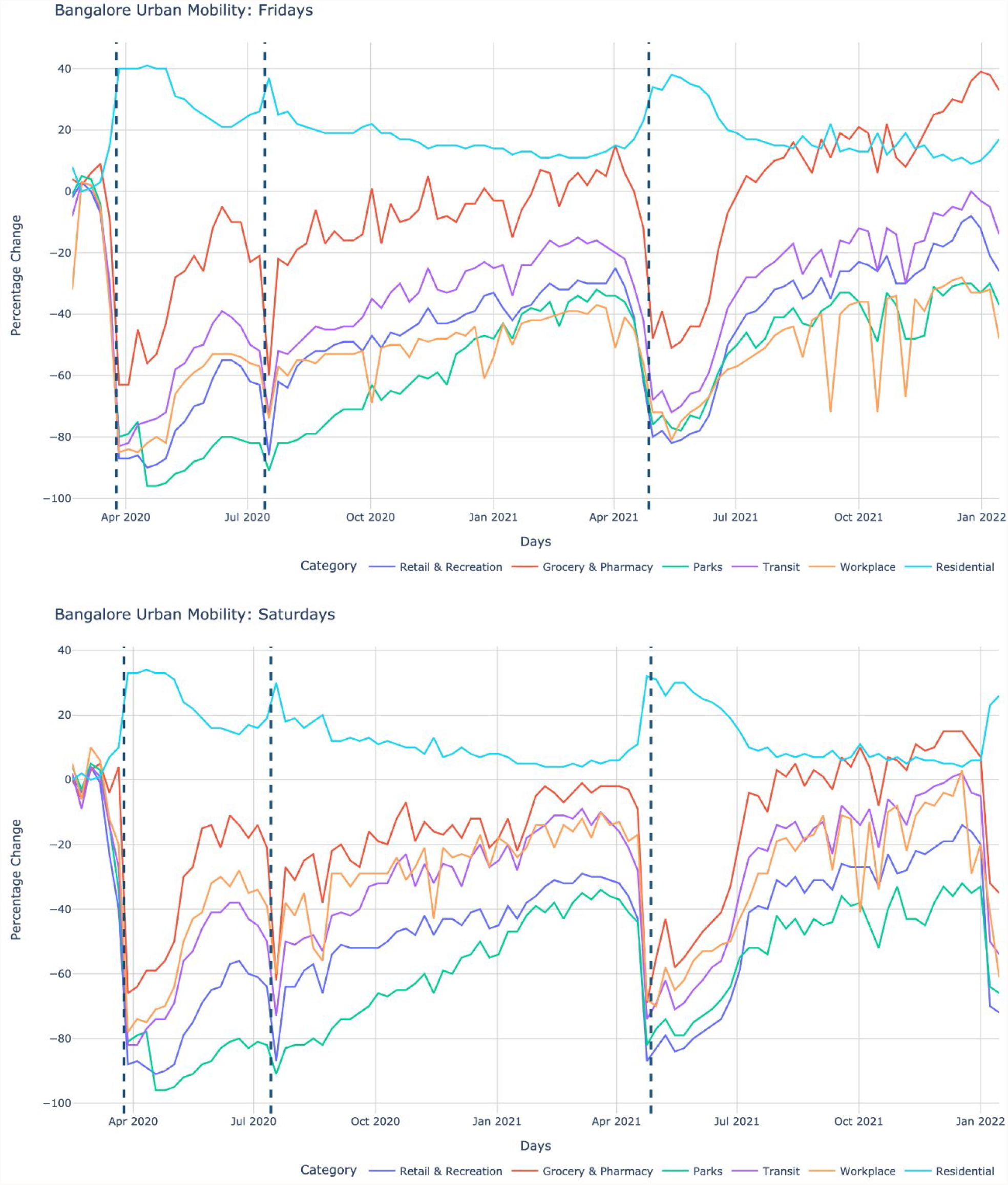
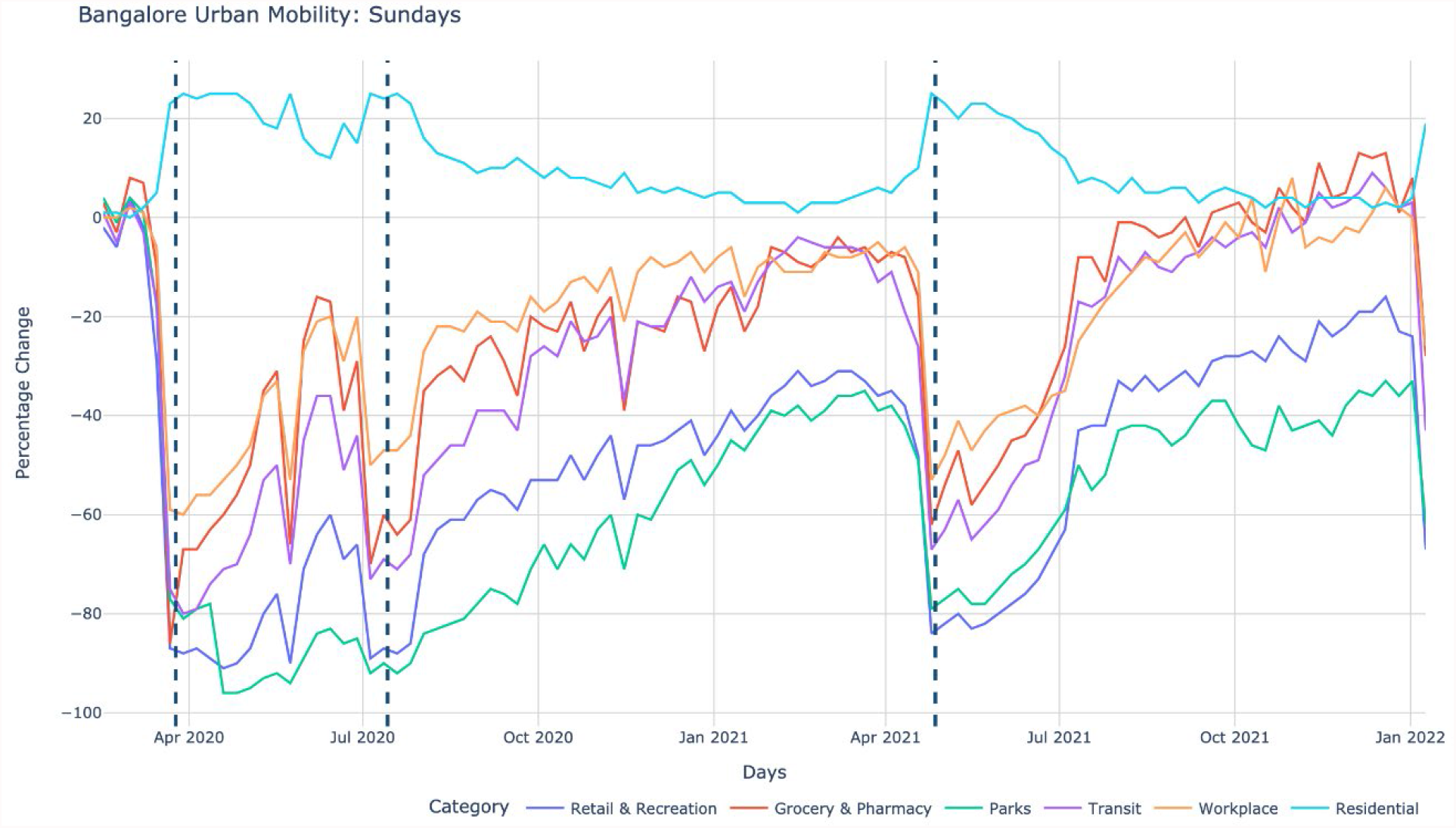
Mobility changes with respect to a baseline pre-pandemic period on each day of the week.

## References

1 Adiga, A. et al., 2021. Strategies to Mitigate COVID-19 Resurgence Assuming Immunity Waning: A Study for Karnataka, India.

2 Bruhat Bengaluru Mahanagara Palike, 2022. BBMP COVID-19 War Room Bulletin. [Online] Available at: https://apps.bbmpgov.in/Covid19/en/media_pdf/Covid_Bengaluru_19_January_2022%20Bulletin-667.pdf [Accessed 23 Jan 2022].

3 Department of Health, Republic of South Africa, 2022. NICD National COVID-19 Hospital Surveillance. [Online] Available at: https://www.nicd.ac.za/wp-content/uploads/2022/01/NICD-COVID-19-Daily-Sentinel-Hospital-Surveillance-report-National-20220102.pdf [Accessed 23 Jan 2022].

4 Google Inc., 2022. COVID-19 Community Mobility Report for Karnataka, India. [Online] Available at: https://www.gstatic.com/covid19/mobility/2022-01-15_IN_Karnataka_Mobility_Report_en-GB.pdf [Accessed 23 Jan 2022].

5 Google Inc., 2022. COVID-19 Community Mobility Reports. [Online] Available at: https://www.google.com/covid19/mobility/

6 ISI-IISc-Team, 2022. Covid-19 States of India and Karnataka District Timeline 20-21. [Online] Available at: https://www.isibang.ac.in/~incovid19/ [Accessed 23 Jan 2022].

7 World Health Organization, 2021. Classification of Omicron (B.1.1.529): SARS-CoV-2 Variant of Concern. [Online] Available at: https://www.who.int/news/item/26-11-2021-classification-of-Omicron-(b.1.1.529)-sars-cov-2-variant-of-concern [Accessed 23 Jan 2022].

